# Behavioural and Social Predictors of COVID-19 Vaccine Uptake among Persons with Disabilities in Kenya

**DOI:** 10.1101/2023.10.03.23296513

**Authors:** Martin Josphat, Rogers Moraro, Jarim Omogi, Abrar Alasmari, Lennah Kanyangi, Rehema Mwema, Sheillah Simiyu, Sarah Kosgei

## Abstract

The uptake of the COVID-19 vaccine by persons with disabilities remains largely unknown in low-and middle-income countries. This evidence gap necessitates disability-focused research to inform improvements in access and inclusion in the last mile of COVID-19 vaccination programs and to support future programs for other vaccine-preventable diseases. We aimed to identify behavioural and social predictors of COVID-19 uptake among persons with disabilities in Kenya. This was a convergent parallel mixed method study that involved questionnaires (792), key informants interviews, and focus group discussions among persons with disabilities and key stakeholders (government actors and professional associations). Data were analysed using STATA statistical analysis software (version 14). Chi-square (*X*^2^) and Fisher’s exact tests were used to test for differences in categorical variables; multivariate regression analysis was employed to ascertain the factors that influence uptake of COVID-19 among persons with disabilities (PWDs) in Kenya. Approximately 59% of persons with disabilities reported to be fully vaccinated, with significant disparities noted among those with cognition (34.2%) and self-care (36.6%) impairments. Confidence in vaccine benefits (Adjusted odds ration [OR]; 11.3, 95% CI; 5.2-24.2), health worker recommendation (OR; 2.6, 95% CI; 1.8-3.7), employment (OR; 2.1, 95% CI; 1.4-3.1), perceived risk (OR; 2.0, 95% CI; 1.3-3.1), age and area of residence were statistically significant predictors of vaccine uptake among PWDs. The primary reasons for low uptake included perceived negative vaccine effects and lack of adequate information. No association was found between having a primary caregiver and/or assistive device, with COVID-19 vaccine uptake. Subsequent vaccination deployments should map and reach PWDs through relevant institutions of PWDs, and localized vaccination campaigns. Related communication strategies should leverage on behaviour change techniques that inspire confidence in vaccines, and on the credibility and trust in health workers to improve vaccine uptake.

## Introduction

Kenya is one of the countries significantly affected by the global pandemic of coronavirus disease 2019, with reported high transmissions resulting in over 0.34 million infected cases and a death toll of over 5,668 [1]. The country’s COVID-19 vaccination uptake however among persons with disabilities (PWDs) in Kenya is unknown, despite the unique barriers they face [2], their special vulnerabilities including chronic conditions, and higher risk for severe outcomes. Through a national COVID-19 vaccine deployment plan, the government of Kenya prioritized vaccination as a key measure to contain COVID-19 spread that also targeted population groups including PWDs scheduled to be vaccinated in phases [3]. However, disaggregated data on proportions of vaccinated PWDs has been conspicuously missing on the COVID-19 vaccination MoH update reports, making it difficult to track the progress made in reaching this cohort [1]. Moreover, despite availability of several studies on drivers of vaccination among other key population groups [4], there is dismal to no evidence on the same among PWDs in low and middle income countries (LMICs).

Approximately 37% of Kenyan adults and 10% of children between ages 12 and 18 had been fully vaccinated as of December 2022 [5]. Willingness for vaccine uptake was reportedly lower among younger people compared to the older, students compared to those working, and in the Coastal and Northeastern region but higher in Nairobi and Rift Valley regions. Moreover, vaccine confidence was significantly associated with vaccine uptake with Kenyans with lower confidence in vaccine safety being more likely to refuse the vaccine [4].Such studies however miss to account for influences specific to PWDs, considering their unique vulnerabilities.

Inadequate information on health inequalities has been identified as a distinct gap in strengthening inclusion and equity in COVID-19 responses. [6] The scanty available evidence globally shows lower COVID-19 vaccination rates compared to those without disability [7] amidst higher risk of severe illness and premature mortality from COVID-19 [8].

We sought to investigate uptake levels of COVID-19 vaccines and determine the demographic, social and behavioural predictors of COVID-19 vaccination among PWDs in Kenya.

## Materials and Methods

### Study Design

We conducted a convergent parallel mixed methods study, in which quantitative and qualitative elements were conducted concurrently in the same phase of the research process, the methods weighed equally, and analysed independently. We employed a cross-sectional study design where both the behaviour and social factors and uptake of COVID-19 interventions were concurrently assessed during the survey.

### Study Setting

The study was conducted in four select Counties in Kenya selected on the basis of their disability prevalence, and by region to represent the main regions in the country to attain a representative rural-urban mix. The counties were namely Embu in Eastern Kenya with the highest disability prevalence of 4.4%, Siaya (4.1%) in the Westerly side, Mombasa (1.4%) in the Coastal region and Nairobi (1.1%) in the central region.

### Participants

Our study participants were PWDs chosen from NCPWDs in the hearing, communication, self-care, cognition, mobility, visual and albinism domains based on the Washington Group on Disability Statistics (WG). Albinism was included as it is classified as a disability in Kenya because of the associated visual impairment and other vulnerabilities in society including mistreatment and exclusion [9].

### Variables

The dependent variable for the study was COVID-19 vaccine uptake, whereas the independent variables included demographics, perceived risk to self, confidence in vaccine benefits, safety, and in health workers, social norms, health worker recommendation, reminder notifications, permission, knowing where to get vaccinated and ease of access.

### Study process, sampling and data collection

Data was collected between 27-31^st^ March 2023, about two years after rolling out of the national COVID-19 vaccination program in Kenya.

PWDs were systematically chosen using a K^th^ of four to reach a sample size of 792 from the National Council for PWDs Registration database within Mombasa, Siaya, Embu and Nairobi counties. The respondents were drawn from men and women above 18 years of age. The respondents were contacted prior to field activity while those not found were replaced by choosing the next participant as per the skipping pattern of four.

A structured questionnaire embedded on a mobile collection tool (KOBO) was used to collect quantitative data from PWDs in the selected counties. Focus Group Discussions (FGDs) were conducted among PWDs and their care givers while Key Informant Interviews (KIIs) were done among disability service officers, social development officers, COVID-19 vaccination in-charges and NCPWDs representatives in the target counties. Behavioral and Social Drivers (BeSD) framework in **Error! Reference source not found.** informed the data collection tools. Research assistants were trained on data collection methods including consenting and interviewing processes, as well as on the various disability domains and related disability etiquette. Interviews were done in languages understandable to the respondents, with sign-language interpreters engaged to administer interviews to the hearing-impaired.

**Fig 1.**
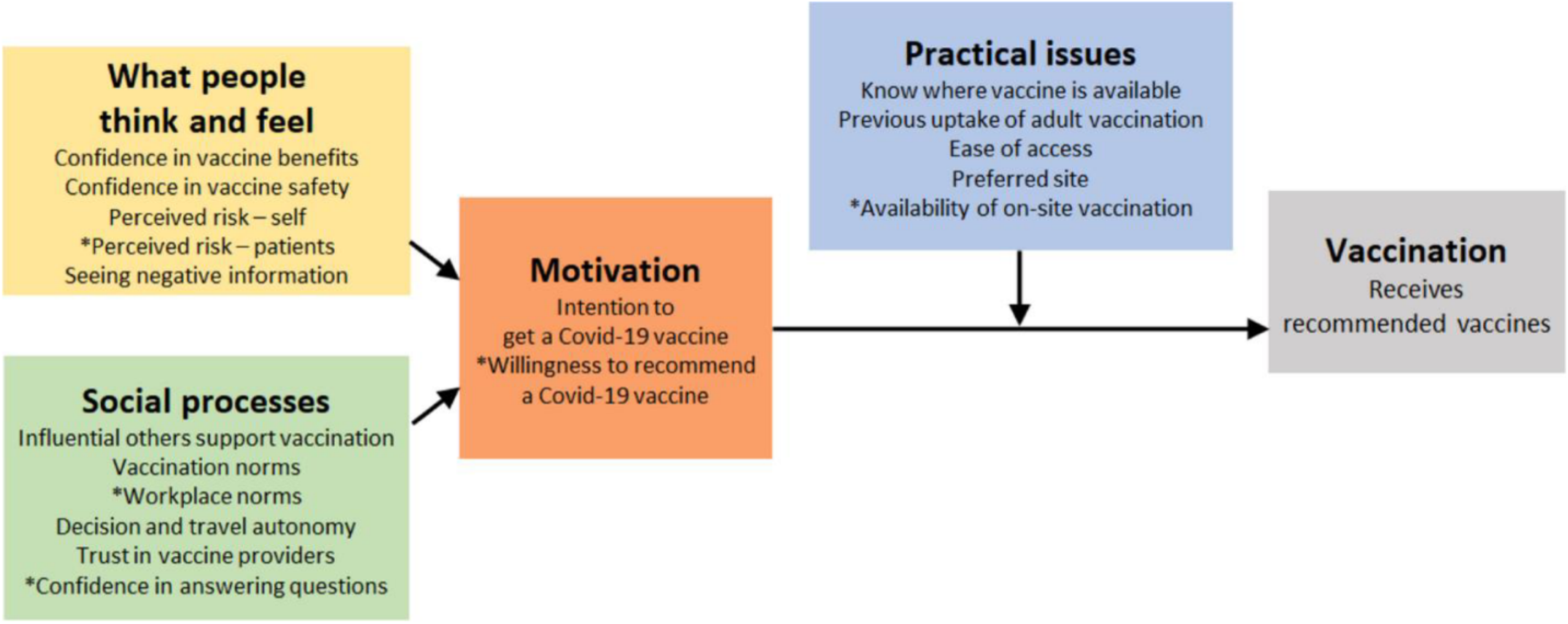
The Behavioral and Social Drivers (BeSD) Framework. Source: Behavioral and social drivers of vaccination: tools and practical guidance for achieving high uptake. Geneva World Health Organization and UNICEF; 2022

### Data Management and Analysis

All data collected through KOBO were first transferred to the Excel for cleaning and later to STATA statistical analysis software (version 14) for analysis.

Descriptive statistics were presented in tables. Univariate analysis was done for all variables to compare outcomes of interest. Proportions were used for categorical variables and measures of central tendency and dispersion for continuous variables. Chi square and fishers test was used to determine significant differences on the key outcomes’ variables. All statistically significant variables were further analyzed using logistic regression and multiple logistic regression to determine the key predictors of vaccine uptake among PWDs.

For qualitative data, all interviews were transcribed and coded for analysis using NVivo software. Data analysis was done using the Behavioral and Social Drivers (BeSD) framework. Deductive content analysis was used for this study. Though the study largely (BeSD) framework to guide the analysis, we further modified it based on findings to develop new themes not covered by the framework but mentioned in interviews. The qualitative data was initially read by (RM) who developed an initial coding framework with input from (JO). This was then shared with (RH) who read through and double coded nine transcripts, selected across the participant’s category to refine the coding framework. With input from other study team members (JA, AA, LK and SK), the differences in the coding framework were then reconciled and coding was done on the themes and sub-themes identified in the final (BeSD) framework. The data was stored in a password protected shared directory on the Amref Health Africa server based on Amref Health Africa ICT data protection policy. All personal identifiers were removed from the dataset prior to archiving in the Amref Health Africa data repository.

### Ethical Approval and consideration

Ethical approval was obtained from Amref Health Africa’s Institutional Review Board (P1383/2023). A written Informed consent was sought from participants who went ahead and signed a copy of the consent form upon being provided with the information about the study and the potential benefits and risks of their involvement.

## Findings

### Univariate Analysis

A total of 792 respondents were interviewed from four counties namely Nairobi 321(40.5%), Embu 188(23.7%), Siaya 166(21%) and Mombasa 117(14.8%). Significant proportions of the PWDs who responded were urban 350(44.4%), and rural dwellers 284(36%), with about 19% residing in peri-urban areas. The mean age of the respondents was 44±0.6 years with more than half being male (56.2%), and most of them being either married (47.6%) or single (41.8%). Majority reported mobility impairments (62.8%) while those with albinism were the least (1.3%). There were averagely 4 persons in the households visited with more than half indicating that they had a primary care giver and 50.9% used an assistive device. A majority (45.8%) had attained secondary education while 35.4% had attained primary education. Slightly more than half (52%) were unemployed with 31% engaged in self-employment activities. Moreover, over 90% were Christians with a paltry being Muslims as shown in Table 1.

**Table 1.** Socio-Demographic characteristics of PWDs participating in the survey in 4 Kenyan counties.

| Variable name | Categories | n (%) |
| --- | --- | --- |
| County | Embu | 188 (23.7) |
|  | Nairobi | 321 (40.5) |
|  | Siaya | 166 (21) |
|  | Mombasa | 117 (14.8) |
| Residence | Rural | 284 (35.9) |
|  | Urban | 350 (44.2) |
|  | Peri-urban | 158 (19.9) |
| Age of participant | Mean (SD) | 44.0 (0.6) |
| Sex | Male | 445 (56.2) |
|  | Female | 346 (43.7) |
|  | Intersex | 1 (0.1) |
| Marital status | Single | 331 (41.8) |
|  | Married | 377 (47.6) |
|  | Others (Divorced, separated or widowed) | 84 (10.6) |
| Disability domain | Mobility | 531 (62.8) |
|  | Seeing | 112 (14.4) |
|  | Cognition | 88 (11.0) |
|  | Hearing | 45 (5.7) |
|  | Self-care | 72 (9.2) |
|  | Communication | 52 (6.7) |
|  | Albinism | 10 (1.3) |
| Number of people in the household | Mean (SD) | 4.5 (0.1) |
| Primary caregiver | Yes | 419 (52.9) |
|  | No | 373 (47.1) |
| Use of assistive devices | Yes | 403 (50.9) |
|  | No | 389 (49.1) |
| Level of Education | Madrasa | 7 (0.9) |
|  | None | 98 (12.4) |
|  | Primary | 280 (35.4) |
|  | Secondary | 363 (45.8) |
|  | Post-secondary | 44 (5.5) |
| Employment Status | Employed | 122 (15.4) |
|  | Farming | 11 (1.4) |
|  | Retired | 9 (1.1) |
|  | Self employed | 246 (31) |
|  | Unemployed | 412 (52) |
| Religion | Christianity | 724 (91.4) |
|  | Islam | 60 (7.6) |
|  | Others (Jewish, atheists, and Buddhists) | 8 (1.0) |

### Bivariate Analysis

Chi-Square test of demographic factors showed that county, residence area, age, marital status, and having certain disabilities notably cognition or selfcare disabilities were significantly associated with vaccine uptake as shown in Table 2. However, there was no significant relationship established between sex or history of COVID-19 diagnosis with vaccine uptake.

**Table 2.**
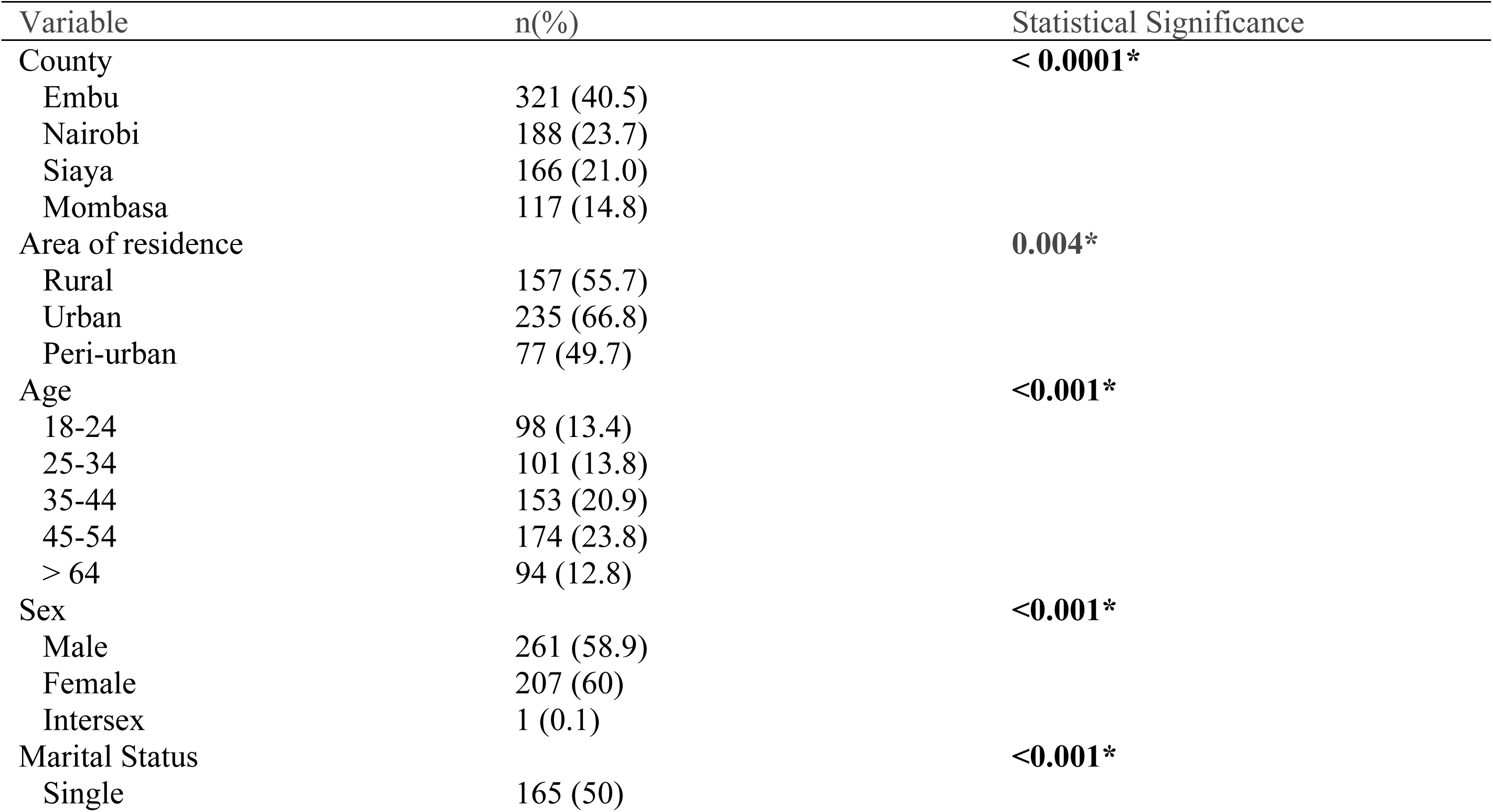
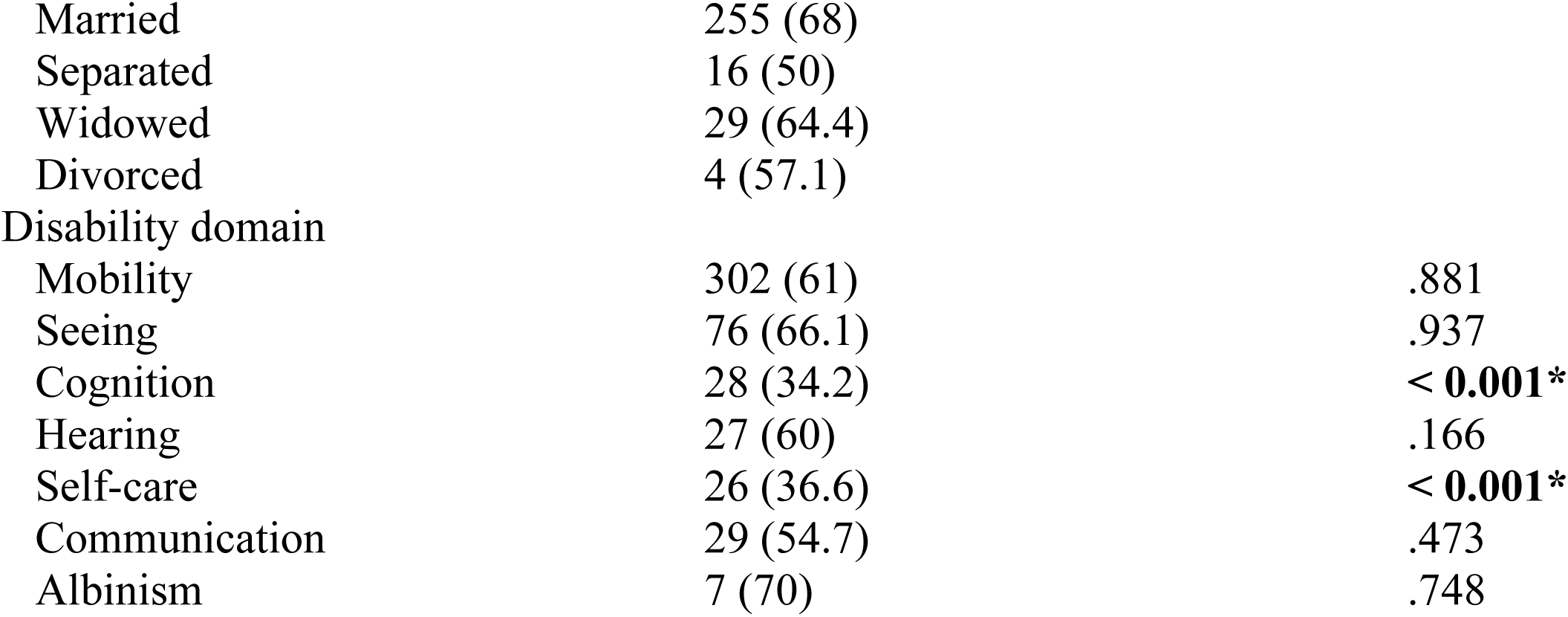
Demographic factors influencing COVID-19 vaccination uptake among PWDs.

| Variable | n(%) | Statistical Significance |
| --- | --- | --- |
| County |  | **< 0.0001\*** |
| Embu | 321 (40.5) |  |
| Nairobi | 188 (23.7) |  |
| Siaya | 166 (21.0) |  |
| Mombasa | 117 (14.8) |  |
| Area of residence |  | **0.004\*** |
| Rural | 157 (55.7) |  |
| Urban | 235 (66.8) |  |
| Peri-urban | 77 (49.7) |  |
| Age |  | **<0.001\*** |
| 18-24 | 98 (13.4) |  |
| 25-34 | 101 (13.8) |  |
| 35-44 | 153 (20.9) |  |
| 45-54 | 174 (23.8) |  |
| > 64 | 94 (12.8) |  |
| Sex |  | **<0.001\*** |
| Male | 261 (58.9) |  |
| Female | 207 (60) |  |
| Intersex | 1 (0.1) |  |
| Marital Status |  | **<0.001\*** |
| Single | 165 (50) |  |
| Married | 255 (68) |  |
| Separated | 16 (50) |  |
| Widowed | 29 (64.4) |  |
| Divorced | 4 (57.1) |  |
| Disability domain |  |  |
| Mobility | 302 (61) | .881 |
| Seeing | 76 (66.1) | .937 |
| Cognition | 28 (34.2) | < **0.001\*** |
| Hearing | 27 (60) | .166 |
| Self-care | 26 (36.6) | < **0.001\*** |
| Communication | 29 (54.7) | .473 |
| Albinism | 7 (70) | .748 |

### Social and Behavioural Factors Influencing Vaccine Uptake

Presence of a primary caregiver, education level, employment status, and religion were associated with vaccine uptake in the univariate analyses, however only employment was statistically significantly associated in the final model. Use of assistive devices was also not associated with vaccine uptake. Individual and societal level factors were shown to influence vaccine uptake including high risk perception, vaccine confidence, family and religious leaders’ norms, recommendation by health worker, recall notification, knowing where to get vaccinated, and ease of access. Whereas community leaders’ opinions not to influence vaccine uptake, qualitative findings reveal an influence of the political class on vaccine uptake. Several accounts were given of getting the vaccine upon seeing their political leaders, notably the president, getting it. No association was also found between one requiring permission to go for the vaccine and getting vaccinated as shown in Table 3.

**Table 3.** Social and behavioural factors influencing COVID-19 vaccination uptake among PWDs.

| Variable | n(%) | Statistical Significance |
| --- | --- | --- |
| Primary care giver |  | 0.003* |
| Yes | 230(54.5) |  |
| No | 239(65.2) |  |
| Use of assistance device |  | 0.864 |
| Yes | 272(67.3) |  |
| No | 197(51.2) |  |
| **Education level** |  | <0.001* |
| None | 34(4.31) |  |
| Madrassa | 3(0.38) |  |
| Primary | 139 (17.6) |  |
| Secondary | 152(19.3) |  |
| Post Secondary | 141 (17.9) |  |
| **Employment Status** |  | <0.001* |
| Formally employed | 102 (79.7) |  |
| Self employed | 167 (69.6) |  |
| Unemployed | 199(48.1) |  |
| **Religion** |  | 0.032* |
| Christianity | 437(60.9) |  |
| Islam | 29(46) |  |
| Others | 3(37.5) |  |
| **Behavioural Factors** |  |  |
| Perceived risk - self | 406(51.1) | <0.001* |
| Confidence in COVID-19 vaccine benefits | 460(64.1) | 0.04 |
| Confidence in vaccine safety | 448(67.1) | <0.03 |
| Confidence in health worker | 752(95) | <0.001* |
| Family Norms | 412(74.9) | <0.001* |
| Religious leader norms | 454(71.4) | <0.001* |
| Community leader norms | 431(69.4) | 0.121 |
| Health worker recommendation | 374(88) | <0.001* |
| Received recall | 334(83.5) | <0.001* |
| Knowledge on where to get vaccinated | 522(96.8) | <0.001* |
| Ease of access | 510(59) | <0.001* |

The key motivations expressed by PWDs for getting vaccinated were to protect themselves (92.9%), protect family (75.5%) and gain access to spaces (30.6%). Qualitative findings further revealed unique motivations to PWDs, for instance getting vaccinated to safeguard oneself against additional exposure from using assistive devices e.g. white cane and reading braille. Low vaccine uptake was mainly attributed to perceived negative vaccine effects (35.3%) and inadequate information (20.5%) with lack of information notably higher among persons with cognition impairments. Perceived vaccine effects were also conspicuously reported among respondents in the qualitative findings, with a majority reporting to have heard of the effects from external sources, and hardly any from personal experience. Other notable reasons were distance and associated cost implications. One of the respondents recounted, *“The vaccination place was far from our residential area. Most of the people didn’t get vaccinated because they didn’t have fare to move from the residential area to where the vaccination was taking place.”* – Visually Impaired FGD Respondent from Siakago, Embu County.

Whereas PWDs believed that their close family & friends and religious leaders wanted them to get vaccinated, they trusted the healthcare workers’ recommendation most (80%), and close family & friends second (39%). Other trusted social contacts were social service officers (14%), caregivers (12%) and religious leaders (8%). The qualitative findings similarly revealed that health workers were considered as credible sources of information and service provider, as recounted for instance: *“If I hear this information from a medical person, I would believe [it] because they have knowledge in the field. I would trust them [healthcare providers], but if I get it [information] from people around here I would not trust the vaccine since people have so many things going on. If it [information] comes from a religious leader, I will accept but not believe since that is not their area of specialty.”* – A Physically Impaired FGD Respondent from Mbeere North, Embu County. Health workers were however required to provide clear information to influence vaccine uptake, as recounted *“If safety is guaranteed and the healthcare providers give clear [information] and sensitize, there will be an increase the uptake.”* – The Regional In-charge of the National Council for PWDs for Meru, Tharaka Nithi, Isiolo and Marsabit counties Despite high trust in health worker, the study found out that over 30% of PWDs had not been reached by health workers and the vaccine recommended to them. Four in ten PWDs reported low ease of access to vaccination services (p < 0.001) with the main reasons cited as difficulty getting to vaccination sites and long waiting times. Notably lower ease of access was reported among persons with selfcare (47.8%), cognition (48.7%) and vision (50%). A paltry (4%) were vaccinated through door-to-door, despite of home administration being the top recommended approach by PWDs on reaching them with vaccines. Other key recommendations made were working with PWD-organizations and their social networks, targeted sensitization of PWDs for informed decisions, special considerations to PWDs, equipping health workers to handle PWDs and/or making transportation arrangement for PWDs to vaccination sites.

### Multivariate Analysis

Further analysis of the statistically significant factors at bivariate level showed age, county, employment status, perceived risk, confidence in vaccine benefits and health worker recommendation were statistically significant predictors of COVID-19 vaccine uptake in our final multiple logistic regression model. According to the findings, the older persons were likely to get vaccinated compared to the younger age groups. For instance, persons aged 35-44 years were two times less likely to be vaccinated compared to those aged 64 years and above.

Respondents from Embu County were less likely to be vaccinated compared to those from Siaya County. The employed according to this data were 2.63 times likely to be vaccinated compared to the unemployed. The findings also show that perceived risk, confidence in the vaccine confidence health worker recommendation are key predictors to vaccine uptake among PWDs as shown in Table 4.

**Table 4.**
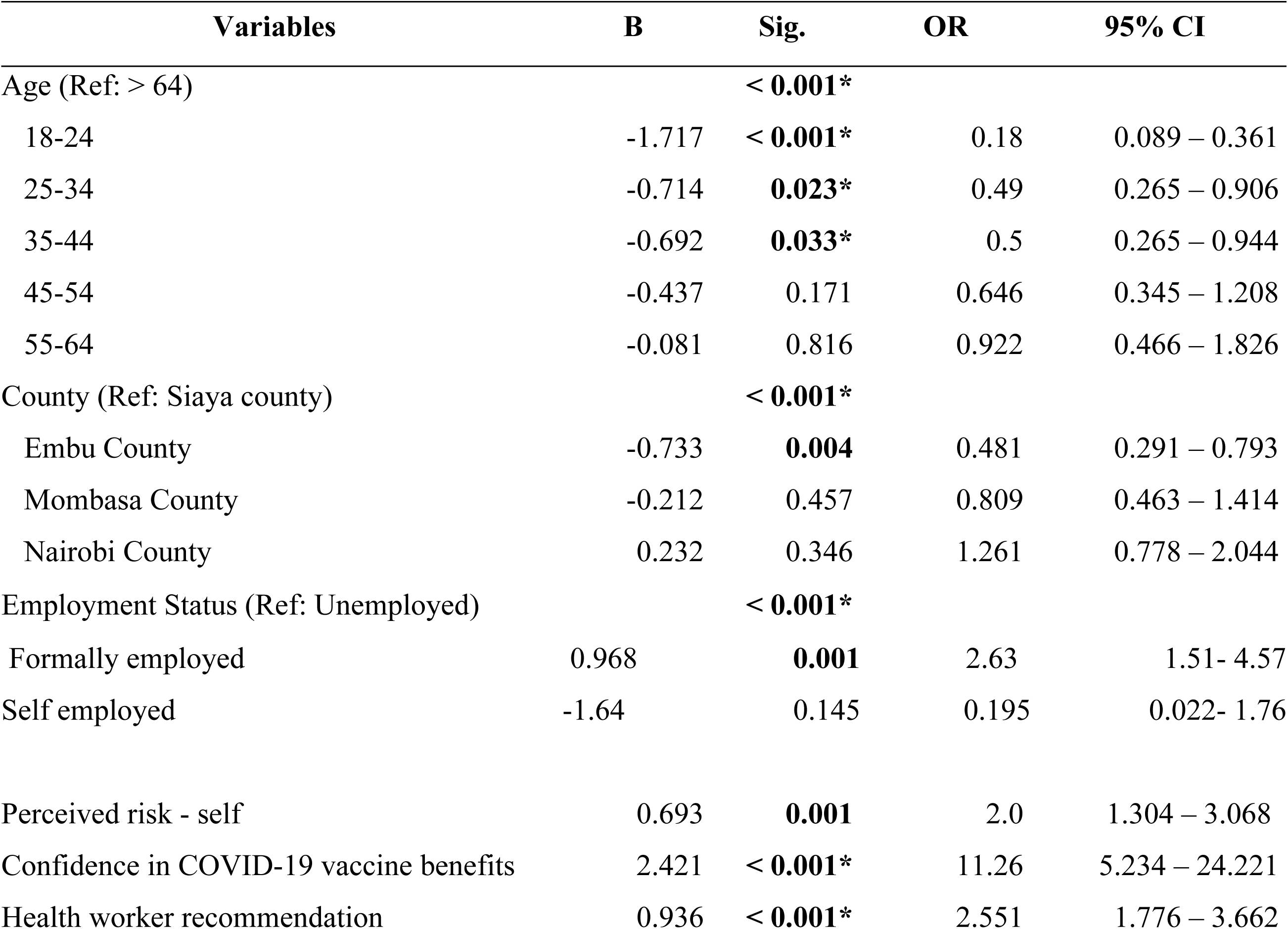
Logistic regression on COVID-19 vaccination uptake predictors among PWDs.

| Variables | B | Sig. | OR | 95% CI |
| --- | --- | --- | --- | --- |
| Age (Ref: > 64) |  | < **0.001** * |  |  |
| 18-24 | -1.717 | < **0.001** * | 0.18 | 0.089 – 0.361 |
| 25-34 | -0.714 | **0.023** * | 0.49 | 0.265 – 0.906 |
| 35-44 | -0.692 | **0.033** * | 0.5 | 0.265 – 0.944 |
| 45-54 | -0.437 | 0.171 | 0.646 | 0.345 – 1.208 |
| 55-64 | -0.081 | 0.816 | 0.922 | 0.466 – 1.826 |
| County (Ref: Siaya county) |  | < **0.001** * |  |  |
| Embu County | -0.733 | **0.004** | 0.481 | 0.291 – 0.793 |
| Mombasa County | -0.212 | 0.457 | 0.809 | 0.463 – 1.414 |
| Nairobi County | 0.232 | 0.346 | 1.261 | 0.778 – 2.044 |
| Employment Status (Ref: Unemployed) |  | < **0.001** * |  |  |
| Formally employed | 0.968 | **0.001** | 2.63 | 1.51- 4.57 |
| Self employed | -1.64 | 0.145 | 0.195 | 0.022- 1.76 |
| Perceived risk - self | 0.693 | **0.001** | 2.0 | 1.304 – 3.068 |
| Confidence in COVID-19 vaccine benefits | 2.421 | < **0.001** * | 11.26 | 5.234 – 24.221 |
| Health worker recommendation | 0.936 | < **0.001** * | 2.551 | 1.776 – 3.662 |

## Discussion

The proportion of PWDs respondents who indicated they had been fully vaccinated (59%) was higher than the general population proportion vaccinated in Kenya (36.9%) within the same period. This points to deliberate interventions to get PWDs vaccinated, and higher acceptance levels. However, those in the self-care and cognition domains were disproportionately vaccinated compared to other domains. This could be partly attributed to the low vaccine confidence and low ease of access among both domains, coupled with lack of information for the cognitively impaired, as established by our study. Selfcare and cognition disabilities have been linked to communication barriers, difficulty understanding information, and dependency on caregivers. A survey by PAHO found that persons with cognitive or intellectual disabilities lacked easy read, simpler text versions of public information and communication material which made decision making difficult. [12] A study among people with intellectual and developmental disabilities and their families showed that COVID-19 vaccine uptake was associated with self-reported knowledge about the vaccine and learning about the vaccine from one’s doctor among other variables. [13] Accessible education and support from healthcare providers and caregivers is significant in addressing these disparities. [2]

As in past studies [14] [15] [16],living in rural was associated with low vaccine uptake, however these finding was not statistically associated with vaccine uptake in our study’s final multiple logistic regression. The study also revealed no statistically significant association between primary caregiver or usage of assistive technology by PWDs with COVID-19 vaccine uptake. The qualitative findings did however show that PWD have special reasons, such as protecting oneself against additional exposure from using supportive devices like a white cane or reading braille. These findings imply that even if the use of assistive devices may not directly affect vaccination rates, PWDs may have particular concerns relating to their condition that affect their vaccination choices. On the low vaccine uptake among the youth, Osur *et al* established the main causes of vaccine hesitancy among youth in Kenya to be concerns about vaccine safety and effectiveness, with social media as the major source of information contributing to hesitancy. [17] Higher vaccine hesitancy was also reported in younger persons with intellectual and developmental disabilities in New York state individuals by [18]. Some studies have also reported lower likelihood of vaccine hesitancy among the elderly [19] [20].Past related studies in Kenya have also shown low confidence in COVID-19 vaccines in the coast region (67%), Nyanza (76%) and Eastern (78%) compared to Nairobi (79%). This aligns with our findings of significantly low vaccine uptake in Mombasa in the coastal region. A study by Orangi *et al* on determinants of vaccine confidence in Kenya however presents contradicting results that rural counties had higher odds of reporting vaccine hesitancy (aOR=2.46; 95% CI: 1.02-5.94) as compared to those in urban counties like Mombasa. [16] However, unlike many studies that show no association between employment and vaccine uptake among other population cohorts, our study’s final result showed a direct significant association among PWDs. The fact that vaccination is required for access to services and for employees by the government may explain the higher vaccination rates among employed PWDs.

On the other hand, costs associated with including transport expenses would have been difficult for those without jobs. Decentralizing vaccination services to PWDs and considerations for free transportation to vaccination facilities may increase vaccine uptake rates among the unemployed PWDs.

The strong influence of high vaccine confidence and risk perception is corroborated in prior studies [21], [22], [23], [24] with the pandemic reported to have had a positive impact on general vaccine confidence in Kenya. ^c^ Masters et al also cited low confidence in COVID-19 vaccine as the strongest correlate of not taking COVID-19 vaccines (adjusted prevalence ratio = 5.19, 95% CI = 4.93 – 5.47) [25]. Orangi et al also linked risk perception to vaccine confidence with those perceiving COVID-19 as not risky having 1.8 times higher odds of being vaccine hesitant compared to those who perceived the disease as risky. [16] High risk perceptions were also attributed to vaccine acceptance (aOR = 2.76, 95% CI: 1.48-5.14) among people living with HIV [26]. However, in a study by Alobaidi S, perceived severity was not significant in predicting vaccine uptake intentions. [27]. Vaccination communication campaigns and advocates should inevitably consider inspiring vaccine confidence. The “Stop HPV, Stop Cervical Cancer” by the Danish Health Authority, the Danish Cancer Society and the Danish Medical Association is a classical campaign building vaccine confidence. [28] The key motivations of protecting oneself as well as significant others, as established in the study, could be capitalized on alongside the messages raising vaccine confidence. While communicating to PWDs however, the motivations should be contextualized to include routes of transmission unique to this cohort including possible infection through assistive devices and caregivers.

The major impediment to vaccine uptake was perceived vaccine effects and has also been cited in past studies [29], [23], [17], [16], [27], even among health workers [30] [31] In a study among American with disabilities, vaccine effects were similarly glaring with the study highlighting higher concerns about COVID-19 vaccine safety as compared to concerns about contracting the disease. Our study revealed further that most of the purported effects were heard from external sources mainly social contacts. This points to the need to provide guidance on credible sources of information to minimize conflicting and misleading information.

Healthcare providers and family members have been similarly cited as trusted sources of information by PWDs. [18]Latkin et al also found out that close family and friends discouraging vaccination was a key predictor of low vaccine uptake (aOR = 0.26, 95% CI = 0.07-0.98). Vaccination programs and influencers including social services officers and religious leaders should therefore not only reach PWDs, but their close social contacts too, considering the influence they hold. The high trust in health workers has been consistently attributed to [32], [33], [34], [35] with the information they provide being linked to better health access [36]. With vaccine hesitancy reported even among health workers [37] despite perceived COVID-19 severity, prevention and vaccine safety, a study by El-Sokkary et al recommends a multidimensional approach to increasing vaccine acceptability [38]. Many healthcare professionals however lack information and may feel hesitant to provide accurate responses [39] which necessitates capacity building of health workers and other influencers of PWDs to adequately address patients’ questions and concerns regarding vaccination. Communication on vaccine uptake should be grounded in the key principles of science-based evidence and data, transparency (i.e. acknowledging to the public what is not yet known), and communicating clearly to achieve understanding by all persons, as recommended by NIH’s Dr. Anthony Fauci. [40].This would help bridge perceived negative effects of vaccines and lack of information that were mainly linked to vaccine hesitancy.

The low vaccine uptake attributed to low ease of access has been shown to be bridged by localizing vaccination services which reduces the difficulty to get to vaccination centers. A study found that a community-based health effort utilizing a mobile vaccination clinic, successfully increased COVID-19 vaccine adherence among the Black population in San Bernardino County with observed lower uptake rates [41]. The desire for house visits and mobile clinics to increase accessibility was overemphasized in our qualitative results too, with higher preference for door-to-door immunization expressed by PWDs. Prioritization of vulnerable populations and centralized fixed-time appointments for receiving the vaccines could potential save them from the long waiting. To improve vaccine uptake among these cohort, working with PWDs’ organizations and networks, focusing awareness efforts, creating custom accommodations and specialized training for healthcare professionals have also been recommended.

### Generalizability

The results of our study can be extrapolated and applied to PWDs outside the sampled population to inform vaccination programs’ decisions targeting the population cohort and their families. The external validity of our study is supported by the consistency of our results with those found in studies in other populations. The studies cross examined were similarly based on study designs with clearly-stated hypotheses in well-defined populations, and on a globally-applied Behavioral and Social Drivers (BeSD) of vaccination framework.

## Conclusion

In conclusion, this study highlights the uneven COVID-19 vaccine uptake across disability domains, with lower rates for cognition and self-care impairments. Our study found out that confidence in vaccine benefits, being employed, perceived self-risk and health worker recommendation were associated with high vaccine uptake, whereas perceived effects were attributed to low uptake. The influence of health worker recommendations on vaccination choices highlights the significance of healthcare provider participation in vaccine outreach. Improving immunization rates among PWDs requires addressing issues with access, vaccination site difficulties, and long waiting times. Targeted tactics including collaborating with disability organizations, raising awareness among PWDs, and setting up transportation can be put into practice to guarantee that everyone has fair access to vaccinations.

## Data Availability

The authors have attached the raw quantitative data

## References

1. MOH. COVID-19. 2022; Available from: https://www.health.go.ke/covid-19.

2. Wiggins, L.D., H. Jett, and J. Meunier, Ensuring equitable COVID-19 vaccination for people with disabilities and their caregivers. Public Health Reports, 2022. 137(2): p. 185–189 DOI: 10.1177/00333549211058733.

3. MOH, National COVID-19 Vaccines Deployment and Vaccination Plan. January 2021. p. 67.

4. CDC, A., COVID-19 Vaccine Perceptions: A 15-country study. February 2021.

5. MOH. MoH (2022) Kenya COVID-19 Vaccination Program – Daily Situation Report. 2022 [cited 2023 24th July]; Available from: Available at: https://www.health.go.ke/covid-19.

6. Pearce, E., et al., Promoting equity in health emergencies through health systems strengthening: lessons learned from disability inclusion in the COVID-19 pandemic. International journal for equity in health, 2022. 21(3): p. 1–7 DOI: 10.1186/s12939-022-01766-6.

7. Wiegand, H.F., et al., COVID-19 vaccination rates in hospitalized mentally ill patients compared to the general population in Germany: Results from the COVID Ψ Vac study. European Psychiatry, 2022. 65(1): p. e41 DOI: 10.1192/j.eurpsy.2022.33.

8. Melo, D.C.F.d., et al., People with Disabilities and COVID-19 in the state of Espírito Santo, Brazil: between invisibility and lack of Public Policies. Ciência & Saúde Coletiva, 2022. 27: p. 4203–4212 DOI: 10.1590/1413-812320222711.08232022EN

9. KNCHR (2022) Mapping Laws, P.a.P.o.A.a.E.G.a.M.O.o.P.w.A.i.K.r.N., Nairobi County: Tab Lights, pp. viii–ix., *MAPPING LAWS*, POLICIES AND PROGRAMMES. 2022.

10. Arina Anis Azlan, M.R.H., Tham Jen Sern, Suffian Hadi Ayub, Emma Mohamad *Public knowledge, attitudes and practices towards COVID-19: A cross-sectional study in Malaysia.* PLoS One, May 21, 2020. 5(15) DOI: 10.1371/journal.pone.0233668.

11. KNBS, Status of disability in Kenya Statistics from the 2019 census. May 2020, Kenya National Bureau of Statistics (KNBS). p. 6–7.

12. PAHO. PAHO pilot program aims to improve Covid-19 vaccine uptake for persons with disability in the Americas. December 3, 2022 [cited 2023 31.08.2023]; Available from: https://www.paho.org/en/stories/paho-pilot-program-aims-improve-covid-19-vaccine-uptake-persons-disability-americas.

13. Lineberry, S., et al., The role of information and knowledge in COVID-19 vaccination among people with intellectual and developmental disabilities and their families. Intellectual and Developmental Disabilities, 2023. 61(1): p. 16–30 DOI: 10.1352/1934-9556-61.1.16.

14. Sun, Y., et al., What attributes influence rural household’s willingness to get vaccinated for COVID-19? Perspectives from six Chinese townships. Vaccine, 2023. 41(3): p. 702–715 DOI: 10.1016/j.vaccine.2022.11.062.

15. Till, B. and T. Niederkrotenthaler, Predictors of vaccine hesitancy during the COVID-19 pandemic in Austria: A population-based cross-sectional study. Wiener klinische Wochenschrift, 2022. 134(23-24): p. 822–827 DOI: 10.1007/s00508-022-02061-8.

16. Orangi, S., et al., Assessing the level and determinants of COVID-19 vaccine confidence in Kenya. Vaccines, 2021. 9(8): p. 936 DOI: 10.3390/vaccines9080936

17. Osur, J.O., et al., Determinants of COVID-19 vaccine behaviour intentions among the youth in Kenya: a cross-sectional study. Archives of Public Health, 2022. 80(1): p. 159 DOI: 10.1186/s13690-022-00904-4.

18. Iadarola, S., et al., COVID-19 vaccine perceptions in New York State’s intellectual and developmental disabilities community. Disability and health journal, 2022. 15(1): p. 101178 DOI: 10.1016/j.dhjo.2021.101178.

19. Al-Mohaithef, M. and B.K. Padhi, Determinants of COVID-19 vaccine acceptance in Saudi Arabia: a web-based national survey. Journal of multidisciplinary healthcare, 2020: p. 1657–1663 DOI: 10.2147/JMDH.S276771.

20. Murphy, J., et al., Psychological characteristics associated with COVID-19 vaccine hesitancy and resistance in Ireland and the United Kingdom. Nature communications, 2021. 12(1): p. 29 DOI: 10.1038/s41467-020-20226-9.

21. Mtei, M., et al., Confidence in COVID-19 vaccine effectiveness and safety and its effect on vaccine uptake in Tanzania: A community-based cross-sectional study. Human Vaccines & Immunotherapeutics, 2023. 19(1): p. 2191576 DOI: 10.1080/21645515.2023.2191576.

22. Hilverda, F. and M. Vollmann, The role of risk perception in students’ COVID-19 vaccine uptake: a longitudinal study. Vaccines, 2021. 10(1): p. 22 DOI: 10.3390/vaccines10010022.

23. Roberts, L.W., et al., Self-reported influences on willingness to receive COVID-19 vaccines among physically ill, mentally ill, and healthy individuals. Journal of Psychiatric Research, 2022. 155: p. 501–510 DOI: 10.1016/j.jpsychires.2022.09.017.

24. Bonner, K.E., et al., Behavioral and social drivers of COVID-19 vaccination in the United States, August–November 2021. American journal of preventive medicine, 2023. 64(6): p. 865–876 DOI: S0749379723000168.

25. Masters, N.B., et al., Geographic heterogeneity in behavioral and social drivers of COVID-19 vaccination. American Journal of Preventive Medicine, 2022. 63(6): p. 883–893 DOI: 10.1016/j.amepre.2022.06.016.

26. Iliyasu, Z., et al., Predictors of COVID-19 vaccine acceptability among patients living with HIV in northern Nigeria: a mixed methods study. Current HIV Research, 2022. 20(1): p. 82–90 DOI: 10.2174/1570162X19666211217093223.

27. Alobaidi, S., Predictors of intent to receive the COVID-19 vaccination among the population in the Kingdom of Saudi Arabia: a survey study. Journal of Multidisciplinary Healthcare, 2021: p. 1119–1128 DOI: 10.2147/JMDH.S306654.

28. WHO. Denmark campaign rebuilds confidence in HPV vaccination. 2023 27 February 2018; Available from: https://www.who.int/news-room/feature-stories/detail/denmark-campaign-rebuilds-confidence-in-hpv-vaccination.

29. Myers, A., C. Ipsen, and A. Lissau, COVID-19 vaccination hesitancy among Americans with disabilities aged 18-65: An exploratory analysis. Disability and Health Journal, 2022. 15(1): p. 101223 DOI: 10.1016/j.dhjo.2021.101223.

30. Shiferie, F., et al., Exploring reasons for COVID-19 vaccine hesitancy among healthcare providers in Ethiopia. Pan African Medical Journal, 2021. 40(1) DOI: 10.11604/pamj.2021.40.213.30699.

31. Mohammed, R., et al., COVID-19 vaccine hesitancy among Ethiopian healthcare workers. PloS one, 2021. 16(12): p. e0261125 DOI: 10.1371/journal.pone.0261125.

32. Figueiras, M.J., et al., Levels of trust in information sources as a predictor of protective health behaviors during COVID-19 pandemic: a UAE cross-sectional study. Frontiers in Psychology, 2021. 12: p. 633550 DOI: 10.3389/fpsyg.2021.633550.

33. Hesse, B.W., et al., Trust and sources of health information: the impact of the Internet and its implications for health care providers: findings from the first Health Information National Trends Survey. Archives of internal medicine, 2005. 165(22): p. 2618–2624 DOI: 10.1001/archinte.165.22.2618.

34. Alduraywish, S.A., et al., Sources of health information and their impacts on medical knowledge perception among the Saudi Arabian population: Cross-sectional study. Journal of Medical Internet Research, 2020. 22(3): p. e14414 DOI: 10.2196/14414.

35. Almaazmi, M.A., et al. The Usage and Trustworthiness of Various Health Information Sources in the United Arab Emirates: An Online National Cross-Sectional Survey. in Healthcare. 2023. MDPI DOI: 10.3390/healthcare11050663.

36. Swoboda, C.M., et al., Odds of talking to healthcare providers as the initial source of healthcare information: updated cross-sectional results from the Health Information National Trends Survey (HINTS). BMC Family Practice, 2018. 19(1): p. 1–9 DOI: 10.1186/s12875-018-0805-7.

37. Iliyasu, Z., et al., ‘why should i take the covid-19 vaccine after recovering from the disease?’a mixed-methods study of correlates of covid-19 vaccine acceptability among health workers in northern nigeria. Pathogens and Global Health, 2022. 116(4): p. 254–262 DOI: 10.1080/20477724.2021.2011674.

38. El-Sokkary, R.H., et al., Predictors of COVID-19 vaccine hesitancy among Egyptian healthcare workers: a cross-sectional study. BMC infectious diseases, 2021. 21: p. 1–9 DOI: 10.1186/s12879-021-06392-1.

39. Alasmari, A., H.J. Larson, and E. Karafillakis, A mixed methods study of health care professionals’ attitudes towards vaccination in 15 countries. Vaccine: X, 2022. 12: p. 100219 DOI: 10.1016/j.jvacx.2022.100219.

40. Allegrante, M.E.A.a.J., Authoritative vs. Credible Sources of COVID Health Information: When Necessary is not Sufficient. 2021.

41. Jacinda C. A-M S.C., Veatrice J., Andrea K., Kelvin S., Michael D. H., Juan C. B., Ricardo P., Jennifer V., A three-tiered approach to address barriers to COVID-19 vaccine delivery in the Black community. the lancet Global Health, 2021. 9(6): p. E749–E750 DOI: 10.1016/S2214-109X(21)00099-1.

